# COVID-19 Mortality Risk Assessment: An International Multi-Center Study

**DOI:** 10.1101/2020.07.07.20148304

**Authors:** Dimitris Bertsimas, Galit Lukin, Luca Mingardi, Omid Nohadani, Agni Orfanoudaki, Bartolomeo Stellato, Holly Wiberg, Sara Gonzalez-Garcia, Carlos Luis Parra-Calderón, The Hellenic COVID-19 Study Group, Kenneth Robinson, Michelle Schneider, Barry Stein, Alberto Estirado, Lia a Beccara, Rosario Canino, Martina Dal Bello, Federica Pezzetti, Angelo Pan

**Author notes:** **Corresponding Author:** Dimitris Bertsimas, Sloan School of Management, Massachusetts Institute of Technology, MA, USA. These authors contributed equally to this work (Luca Mingardi, Agni Orfanoudaki, Bartolomeo Stellato, Holly Wiberg).

## Abstract

**Background:** Timely identification of COVID-19 patients at high risk of mortality can significantly improve patient management and resource allocation within hospitals. This study seeks to develop and validate a data-driven personalized mortality risk calculator for hospitalized COVID-19 patients.

**Methods:** De-identified data was obtained for 3,927 COVID-19 positive patients from six independent centers, comprising 33 different hospitals. Demographic, clinical, and laboratory variables were collected at hospital admission. The COVID-19 Mortality Risk (CMR) tool was developed using the XGBoost algorithm to predict mortality. Its discrimination performance was subsequently evaluated on three validation cohorts.

**Findings:** The derivation cohort of 3,062 patients has an observed mortality rate of 26.84%. Increased age, decreased oxygen saturation (≤ 93%), elevated levels of C-reactive protein (≥ 130 mg/L), blood urea nitrogen (≥ 18 mg/dL), and blood creatinine (≥ 1.2 mg/dL) were identified as primary risk factors, validating clinical findings. The model obtains out-of-sample AUCs of 0.90 (95% CI, 0.87-0.94) on the derivation cohort. In the validation cohorts, the model obtains AUCs of 0.92 (95% CI, 0.88-0.95) on Seville patients, 0.87 (95% CI, 0.84-0.91) on Hellenic COVID-19 Study Group patients, and 0.81 (95% CI, 0.76-0.85) on Hartford Hospital patients. The CMR tool is available as an online application at covidanalytics.io/mortality_calculator and is currently in clinical use.

**Interpretation:** The CMR model leverages machine learning to generate accurate mortality predictions using commonly available clinical features. This is the first risk score trained and validated on a cohort of COVID-19 patients from Europe and the United States.

**Research in context:** *Evidence before this study:* We searched PubMed, BioRxiv, MedRxiv, arXiv, and SSRN for peer-reviewed articles, preprints, and research reports in English from inception to March 25^th^, 2020 focusing on disease severity and mortality risk scores for patients that had been infected with severe acute respiratory syndrome coronavirus 2 (SARS-CoV-2). Earlier investigations showed promise at predicting COVID-19 disease severity using data at admission. However, existing work was limited by its data scope, either relying on a single center with rich clinical information or broader cohort with sparse clinical information. No analysis has leveraged Electronic Health Records data from an international multi-center cohort from both Europe and the United States.

*Added value of this study:* We present the first multi-center COVID-19 mortality risk study that uses Electronic Health Records data from 3,062 patients across four different countries, including Greece, Italy, Spain, and the United States, encompassing 33 hospitals. We employed state-of-the-art machine learning techniques to develop a personalized COVID-19 mortality risk (CMR) score for hospitalized patients upon admission based on clinical features including vitals, lab results, and comorbidities. The model validates clinical findings of mortality risk factors and exhibits strong performance, with AUCs ranging from 0.81 to 0.92 across external validation cohorts. The model identifies increased age as a primary mortality predictor, consistent with observed disease trends and subsequent public health guidelines. Additionally, among the vital and lab values collected at admission, decreased oxygen saturation (≤ 93%) and elevated levels of C-reactive protein (≥ 130 mg/L), blood urea nitrogen (≥ 18 mg/dL), blood creatinine (≥ 1.2 mg/dL), and blood glucose (≥180 mg/dL) are highlighted as key biomarkers of mortality risk. These findings corroborate previous studies that link COVID-19 severity to hypoxemia, impaired kidney function, and diabetes. These features are also consistent with risk factors used in severity risk scores for related respiratory conditions such as community-acquired pneumonia.

*Implications of all the available evidence:* Our work presents the development and validation of a personalized mortality risk score. We take a data-driven approach to derive insights from Electronic Health Records data spanning Europe and the United States. While many existing papers on COVID-19 clinical characteristics and risk factors are based on Chinese hospital data, the similarities in our findings suggest consistency in the disease characteristics across international cohorts. Additionally, our machine learning model offers a novel approach to understanding the disease and its risk factors. By creating a single comprehensive risk score that integrates various admission data components, the calculator offers a streamlined way of evaluating COVID-19 patients upon admission to augment clinical expertise. The CMR model provides a valuable clinical decision support tool for patient triage and care management, improving risk estimation early within admission, that can significantly affect the daily practice of physicians.

## Introduction

The ongoing coronavirus disease pandemic (COVID-19) caused by severe acute respiratory syndrome coronavirus 2 (SARS-CoV-2) has led to an alarming number of casualties across the world.^1^ As the pandemic progresses globally, much remains unknown about the disease dynamics and risk factors. A better understanding of the clinical determinants of disease severity can improve patient management throughout the healthcare system. This task is challenging due to the rapid spread of the disease and the lack of detailed patient data.

Leveraging machine learning (ML) methods enables rapid and unbiased insights across large populations of heterogeneous patients. An algorithmic approach provides an objective evaluation and can often capture nonlinear interactions that are not obvious from pure observation of the population. Researchers have recognized the potential of these data-driven approaches across various facets of the effort to combat COVID-19.^2^

In this work, we present the COVID-19 Mortality Risk (CMR) tool, a novel ML model for predicting mortality in hospitalized COVID-19 patients. It enables physicians to better triage patient care in a resource-constrained system through a personalized mortality risk score. The CMR model synthesizes various clinical data elements from multiple European and US centers, including demographics, lab test results, symptoms, and comorbidities. We use the XGBoost algorithm,^3^ a leading ML method, to predict mortality probabilities. This score is able to capture nonlinearities in risk factors, resulting in strong predictive performance with an out-of-sample area under the receiver operating characteristic curve (AUC) of 0.90 (95% CI, 0.87-0.94). It also validates commonly accepted risk factors, such as age and oxygen saturation, while discerning novel insights.

The CMR tool leverages an international cohort from three hospital systems in Italy, Spain, and the United States. The model is subsequently validated on hospitalized patients in a consortium of six hospitals from Greece, Spain, and the United States. Each region presents a diverse set of patient profiles and mortality rates for the model. By considering severely ill populations from different countries and healthcare systems, the final dataset captures a wide array of features.

In recent months, ML scores have been proposed to predict COVID-19 mortality^4,5^ as well as disease severity.^6^ Existing literature largely focuses on Chinese hospitals due to the disease’s emergence in Wuhan.^5,6^ However, it is instrumental to understand the clinical characteristics for more recent and diverse cases, considering that the virus strain may have mutated since surfacing in Wuhan.^7^ Pourhomayoun et al. (2020) proposed a model based on a large international dataset, yet this model lacks comprehensive patient data and is thus limited in its ability to derive personalized insights.^4^ In this work, we study patients in Europe and the US, offering a new lens into the clinical characteristics of this disease.

## Methods

### Study Population

The study comprises 33 different hospitals, spanning across three countries in southern Europe as well as the US. The collaborating institutions were split into derivation and validation cohorts, as summarized in **Table 1**. The derivation cohort includes the healthcare systems of ASST Cremona (Northern Italy), HM Hospitals (Spain), and Hartford HealthCare affiliate hospitals (United States). The broad geographic spread of data sources offers a comprehensive sample of some of the most severely impacted regions in the world. To further validate the results, we partnered with Hospital Universitario Virgen del Rocío (Spain), the Hellenic COVID-19 Study Group (Hellenic CSG), a consortium of Greek hospitals, and Hartford HealthCare’s main hospital (CT, USA). The study population consists of adult patients who were admitted to the hospital with confirmed SARS-CoV-2 infection by polymerase chain reaction testing of nasopharyngeal samples. The time horizon of admissions is displayed in **Table 1**.

**Table 1:** Overview of participating institutions in the derivation and validation cohorts.

| Organization | Region | Study Dates | Hospital Count | Description |
| --- | --- | --- | --- | --- |
| Derivation Cohort |  |  |  |  |
| ASST Cremona | Lombardy (Italy) | 02/01 - 05/08 | 3 | Azienda Socio-Sanitaria Territoriale di Cremona (ASST Cremona) includes the Ospedale di Cremona, Ospedale Oglio Po and other minor public hospitals in the Province of Cremona. Cremona is one of the most hit Italian provinces in Lombardy in the Italian COVID-19 crisis with a total of 4,422 positive cases to date. Ospedale di Cremona has around 750 beds. During the COVID-19 crisis all elective activities and surgeries were suspended and most of the hospital was converted to treat COVID-19. |
| HM Hospitals | Madrid, Galicia, León, Cataluña (Spain) | 02/01 - 04/20 | 17 | HM Hospitals, a leading Hospital Group in Spain with 15 general hospitals and 21 clinical centers that cover the regions of Madrid, Galicia, and León. The group has served more than 2,300 COVID-19 patients over the last two months. Its total capacity includes more than 1,468 beds and 101 operating rooms. |
| Hartford HealthCare (Affiliates) | Connecticut (USA) | 03/18 - 05/14 | 5 | Hartford HealthCare is a major hospital network serving patients throughout Connecticut. In addition to its primary hospital in Hartford, it operates five acute care hospitals: Backus Hospital, Charlotte Hungerford, the Hospital of Central Connecticut, MidState Medical Center, and Windham Hospital. These sites have a total of 1,087 beds and nearly 2,500 physicians on staff. |
| Validation Cohort |  |  |  |  |
| Hospital Universitario Virgen del Rocío | Seville, Andalusia (Spain) | 03/11 - 05/05 | 1 | The Hospital serves a basic population of 557,576 users, between the districts of Seville, Aljarafe and Seville South in the region of Andalusia. It has a provision of 1,279 beds installed, with a staff of 8,409 professionals. During the COVID-19 crisis, elective activities and surgeries were suspended, and most of the hospital was converted to care, COVID-19 patients, it has attended approximately 320 COVID-19 positive cases discharges (exits included) until May 5th. |
| Hellenic COVID-19 Study Consortium | Attika, Thraki, Thessaly, Peloponnese (Greece) | 03/01 - 05/15 | 6 | This is a collection of the referral center of Greece for the management of COVID-19 patients. It includes the Sotiria Thoracic Diseases Hospital of Athens, Evangelismos Hospital, the University Hospitals of Alexandroupolis and Patra, the Attikon General Hospital and the General University Hospital of Larissa. All organizations are independent public (NHS and academic) institutions. |
| Hartford HealthCare (Main Hospital) | Connecticut (USA) | 03/18 - 05/14 | 1 | Hartford Hospital is an 867-bed acute care teaching hospital located in Hartford, Connecticut. Hartford Hospital was established in 1854 and is the central campus of the broader Hartford HealthCare organization. It employs 1,200 physicians and dentists and has a caseload of over 100,000 annual emergency room visits. |

All independent organizations and the Massachusetts Institute of Technology institutional review boards approved this protocol as minimal-risk research using data collected for standard clinical practice and waived the requirement for informed consent. The survey was anonymous and confidentiality of information was assured.

### Clinical Features

Data is collected using the electronic health record databases and COVID-19 specific registries of the collaborating hospitals. We compile 22 features, including patient demographic information, comorbidities, vitals upon admission, and laboratory test results. The full set of features is outlined in **Table 2**. The outcome of interest, mortality during the hospital admission, is derived from discharge records. Only the first recorded laboratory test results are considered, typically within 24 hours of admission. Comorbidities are identified using the International Classification of Diseases, 9th and 10th revision, codes of hospital discharges and are aggregated into four categories using the Clinical Classifications Software.^8^ Missing values are imputed using *k*-nearest neighbors imputation^9^ (details in the Supplementary Material). We exclude risk factors that are not consistently recorded in the derivation cohort, thereby omitting features whose values are more than 40% missing.

**Table 2:** Summary statistics of all patient characteristics for the total sample, the survivor, and non-survivor cohorts.

|  | Median (IQR) |  |  |  |
| --- | --- | --- | --- | --- |
| Filter | All<br>(N = 2,831) | Survivor<br>(N = 2,120) | Non-Survivor<br>(N = 711) | P-Value<sup>a</sup> |
| Age | 68.0 (57.0-79.0) | 63.0 (54.0-74.0) | 81.0 (73.2-86.0) | <1.0E-4 |
| Female* | 1095.0 (38.7%) | 868.0 (40.9%) | 227.0 (31.9%) | <1.0E-4 |
| Heart Rate (bpm) | 89.0 (79.0-101.0) | 90.0 (80.0-102.0) | 87.0 (78.0-100.0) | 1.3E-03 |
| Oxygen Saturation (%) | 94.0 (90.0-96.0) | 94.4 (92.0-96.0) | 88.5 (80.0-93.6) | <1.0E-4 |
| Temperature (°C) | 36.9 (36.4-37.6) | 36.9 (36.4-37.5) | 37.1 (36.5-37.8) | 2.4E-04 |
| Alanine Aminotransferase (U/L) | 27.0 (17.0-44.0) | 27.8 (17.5-45.0) | 25.5 (16.0-41.0) | 3.8E-02 |
| Aspartate Aminotransferase (U/L) | 36.0 (25.0-55.0) | 34.0 (24.4-51.0) | 45.0 (30.0-69.0) | <1.0E-4 |
| Blood Glucose (mg/dL) | 118.0 (105.0-141.0) | 115.0 (103.4-133.0) | 134.0 (113.0-171.0) | <1.0E-4 |
| Blood Urea Nitrogen (mg/dL) | 17.0 (12.6-25.2) | 15.0 (11.5-20.0) | 29.5 (20.3-47.2) | <1.0E-4 |
| C-Reactive Protein (mg/dL) | 74.2 (29.1-149.5) | 58.6 (22.7-119.3) | 141.1 (72.0-223.1) | <1.0E-4 |
| Creatinine (mg/dL) | 1.0 (0.8-1.2) | 0.9 (0.7-1.1) | 1.3 (1.0-1.8) | <1.0E-4 |
| Hemoglobin (U/g) | 13.9 (12.7-15.0) | 14.0 (12.9-15.0) | 13.5 (12.0-14.7) | <1.0E-4 |
| Mean Corpuscular Volume (µm<sup>3</sup>) | 87.8 (84.9-91.0) | 87.5 (84.7-90.4) | 89.3 (85.8-92.7) | <1.0E-4 |
| Platelets (10<sup>3</sup>/µL) | 201.0 (156.0-263.0) | 206.0 (160.0-266.5) | 185.0 (141.0-246.8) | <1.0E-4 |
| Potassium (mEq/L) | 4.1 (3.7-4.4) | 4.0 (3.7-4.4) | 4.1 (3.7-4.6) | 1.4E-04 |
| Prothrombin Time (INR) | 1.1 (1.0-1.2) | 1.1 (1.0-1.2) | 1.1 (1.0-1.3) | <1.0E-4 |
| Sodium (mEq/L) | 137.1 (135.0-140.0) | 137.0 (135.0-139.4) | 138.0 (135.0-141.0) | <1.0E-4 |
| White Blood Cell Count (/µL) | 6800 (5200-9200) | 6500 (5000-8700) | 8000 (5700-11400) | <1.0E-4 |
| Cardiac dysrhythmias* | 200.0 (7.1%) | 127.0 (6.0%) | 73.0 (10.3%) | 6.5E-04 |
| Chronic kidney disease* | 65.0 (2.3%) | 33.0 (1.6%) | 32.0 (4.5%) | 3.7E-04 |
| Heart disease* | 125.0 (4.4%) | 80.0 (3.8%) | 45.0 (6.3%) | 1.1E-02 |
| Diabetes* | 345.0 (12.2%) | 234.0 (11.0%) | 111.0 (15.6%) | 2.7E-03 |

\*Count (proportion) is reported for binary variables.
<sup>a</sup> P-value reports significance of a two-sided T-test between the survivor and non-survivor populations.

### Modeling Approach

We train a binary classification model in which the outcome is patient mortality: 1, if the patient was deceased, or 0, if discharged. Specifically, we use the XGBoost algorithm^4^ for the training process. For comparison, we also present the predictive performance of other ML methods in the Supplementary Material. The derivation population is randomly divided into training (85%) and testing (15%) sets, ensuring that mortality prevalence was consistent between the two. We tune seven model parameters by maximizing the *K*-fold cross-validation AUC using the Optuna optimization framework^10^ (details in the Supplementary Material). This technique provides a more accurate parameter search compared to grid search by efficiently pruning suboptimal parameter combinations and continuously refining the search space. We apply SHapley Additive exPlanations (SHAP) to generate importance plots for transparency of the model predictions and risk drivers.^11^ All statistical analysis is conducted using version 3.7 of the Python programming language.

### Performance Evaluation

All predictive models are evaluated based on their ability to discriminate between outcomes for each population. We report results for the training and testing sets of the derivation cohort, as well as for each independent institution in the validation cohort, with the corresponding confidence intervals (CI). The AUC, accuracy, specificity, precision, and negative predictive value are computed for all patient subpopulations across different thresholds. Receiver operating characteristic (ROC) curves were created for each of the cohorts.

## Results

### Patient Characteristics

The CMR model is created using a derivation population of 3,062 patients, of which 1,441 are from ASST Cremona, 1,390 from HM Hospitals, and 231 from Hartford Affiliates (**Table 2)**. The average observed mortality rate in this population is 26.84%. In comparison to survivors, non-survivors tend to be older (median age 80 vs. 64) and more commonly men (67.2% vs. 58.4% of cohort). Moreover, the prevalence of comorbidities such as cardiac dysrhythmias, chronic kidney disease, and diabetes is higher in the non-survivor population (9.61%, 4.21% and 15.92% versus 5.56%, 1.74%, and 11.42%, respectively); see **Table 2**.

### Performance Metrics

The final mortality model exhibits an out-of-sample AUC of 0.90 (95% CI, 0.87-0.94) on the derivation testing set; see **Table 3**. The AUC for the Seville cohort is slightly higher at 0.92 (95% CI, 0.88-0.95). For the other two validation centers, there is a decrease in AUC. In the Hellenic CSG cohort, the model performs 0.87 (95% CI, 0.84-0.91) and in the Hartford Hospital population 0.81 (95% CI, 0.76-0.85). The corresponding ROC curves are included in the Supplementary Material.

**Table 3.**
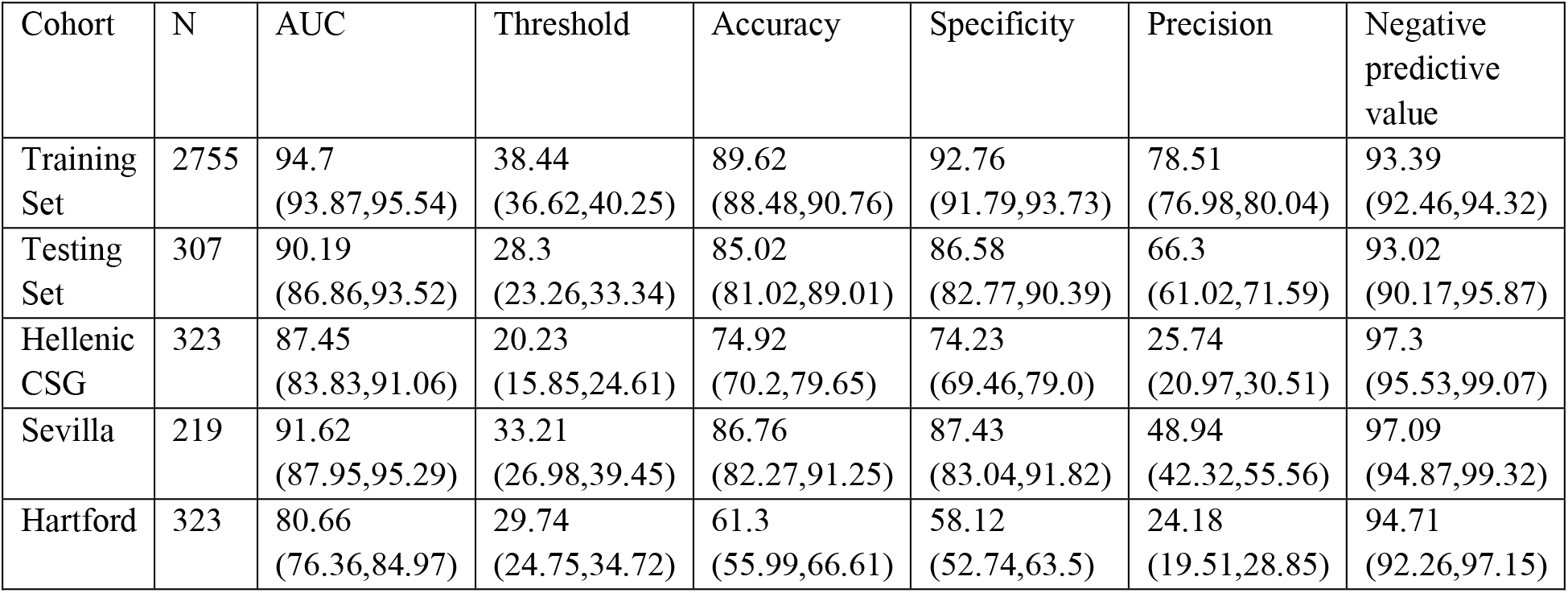
AUC performance (%) and threshold-based metrics for training, testing, and validation populations.

A different threshold is selected for each cohort to enforce a minimum sensitivity of 80%. Given the implications of these predictions, we report conservative risk estimates in order to ensure that all critically ill patients are accounted for. This comes at the expense of specificity, i.e., it increases the number of patients whom we may incorrectly flag as high risk of mortality. For the fixed sensitivity requirement, we achieve a classification accuracy of 0.85 (95% CI, 0.81-0.89) in the testing set with specificity of 0.87 (95% CI, 0.83-0.90); see **Table 3**.

The model generalizes better in the Seville cohort with an accuracy of 0.87 (95% CI, 0.82-0.91) and specificity of 0.87 (95% CI, 0.83-0.92). The necessary threshold for a sensitivity of 80% is lower for the Hellenic CSG compared to the other populations. This is due to the low baseline incidence of mortality in this sample when compared to the derivation and other validation cohorts. The model achieves lower performance in this set of patients, with an accuracy of 0.75 (95% CI, 0.7-0.8) and specificity of 0.74 (95% CI, 0.69-0.79). For Hartford Hospital, the accuracy of CMR is 0.61 (95% CI, 0.56-0.67) with a specificity of 0.58 (95% CI, 0.53-0.64).

### Model Results

Through the SHAP framework, we identified the most important drivers of mortality risk and the interplay between individual features. For a particular patient, SHAP values indicate the feature contributions towards the risk. The patient risk normalized between 0 and 1 is the sum of the SHAP values of all the features (details in the Supplementary Material). **Figure 1(a**) displays the risk contributions of the 10 most important features. For example, higher values of age (red) yield higher SHAP values, suggesting that older patients are at higher risk. In contrast, the SHAP value increases with lower values (blue) of Oxygen Saturation, suggesting an inverse relationship with this feature.

**Figure 1.**
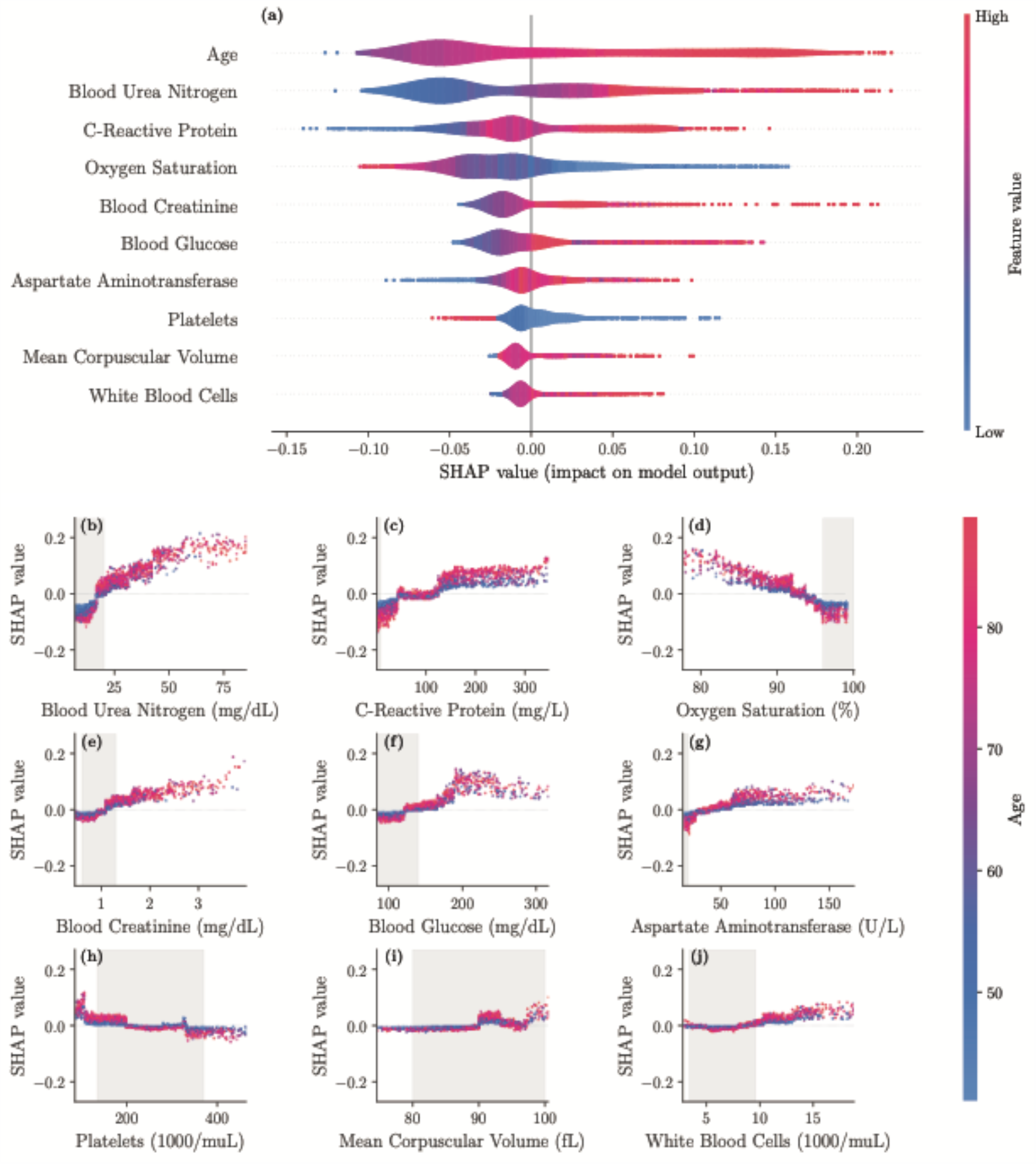
SHAP importance plots for final model. The top 10 features are displayed in panel **a**, ordered by decreasing significance. For a given feature, the corresponding row indicates the SHAP values as the feature ranges from its lowest (blue) to highest (red) value. **Panel b-j** display the individual feature plots and the impact of each feature on the mortality risk (colors indicate the age here) with gray areas indicating reference ranges.

When BUN is below 20 mg/dL, the mortality risk decreases, particularly for ages below 55 years. On the other hand, BUN values greater than 25 mg/dL for older patients increase the risk (**Figure 1(b)**). A C-reactive protein (CRP) between 50 and 130 mg/L does not affect the risk, independent of age. As CRP goes below 50 mg/L, the mortality risk decreases. For a CRP above 160 mg/L, the elevated risk does not change and is higher for older patients (**Figure 1(c)**). An oxygen saturation below 93% increases the mortality risk rapidly and this trend is accelerated by growing age (**Figure 1(d))**. A blood creatinine level greater than 1.2 mg/dL increases the risk moderately, specifically for older patients. Levels above 3 mg/dL rapidly escalate the mortality risk (**Figure 1(e)). Figure 1(f)** illustrates that while a blood glucose less than 130 mg/dL lowers the risk, it can increase the risk for levels above 180 mg/dL, in particular for older patients. An aspartate aminotransferase (AST) level above 65 U/L increases the risk, while a level below 25 U/L sharply decreases the risk, independent of age (**Figure 1(g)**). A platelet count in 10^3^/µL affects the risks in 4 distinct ranges: (i) below 50 the risk is elevated, (ii) between 50 and 180 the risk is marginally increased (more for older patients), (iii) between 180 and 330 the risk is slightly decreased, and (iv) above 330 the risk is sizably decreased (**Figure 1(h)**). **Figure 1(i)** shows that a mean corpuscular volume (MCV) between 90 and 94 fL increases the risk moderately, while other values have only small effects. Lastly, an increased risk is observed when white blood cell (WBC) count is above 10 in 10^3^/µL, in particular for older patients (**Figure 1(j)**).

## Discussion

The CMR calculator predicts mortality with high accuracy using clinical measurements collected early within a patient’s hospital admission. An early risk assessment of patient mortality allows physicians to triage patients and prioritize resources in a highly congested system. It uses commonly available laboratory results and does not require imaging results or advanced testing. The presented tool can be particularly useful in lower acuity facilities or remote hospitals with constrained diagnostic capabilities.

Age is the most important determinant of mortality in the model: older patients have higher mortality risk, which has been observed in retrospective patient analysis^12^ and subsequently reflected in public health guidance.^13^ Predicted mortality also increases for patients with low oxygen saturation, corroborating findings that link hypoxemia to mortality,^14^ as well as the observed prevalence of shortness of breath in severe patients.^15^ This measurement additionally serves as signal of respiratory distress, and respiratory failure has been found clinically as one of the major mortality causes of COVID-19.^16^ This can also appear in cases of silent hypoxia where shortness of breath is not observed.^17^

Our study finds that elevated BUN, CRP, creatinine, glucose, AST, and platelet counts are highly significant laboratory features. Several of these biomarkers have been identified in other retrospective analyses of mortality outcomes of COVID-19.^16,18^ Prior work has also uncovered the critical role of these biomarkers in identifying severe cases of patients with community acquired pneumonia.^19^ The PSI, CURB-65, and SCAP scores are also based on similar risk factors such as glucose levels ≥ 250 mg/dL and BUN > 19 mg/dL.^20,21^ Moreover, CRP levels have been recognized to characterize severity for H1N1 patients.^22^

CRP is a widely available inflammatory marker which has been independently observed as a biomarker of COVID-19 severity.^6,23^ Our findings show that CRP values outside the reference ranges do not necessarily increase the risk of mortality. In fact, CRP has a negative effect on mortality until approximately 50 mg/L, it has a negligible effect between approximately 50mg/L and 130 mg/L, and it significantly increases the mortality risk above 130mg/L. Elevated BUN and creatinine levels are both indicative of impaired kidney function, which has been associated with poor prognosis.^24^ The individual feature plots indicate a clear transition from low to high risk when BUN exceeds approximately 18 mg/dL and creatinine exceeds approximately 1.2 mg/dL. These values are slightly lower than reference ranges for these values, providing data-driven validation of the ranges^25^ targeted for COVID-19. The increase in mortality risk for patients with elevated glucose levels is consistent with the reports in other studies of diabetes as a risk factor.^12,26^ Elevated AST levels have been observed due to liver dysfunction in severe COVID-19 cases.^27^ Finally, low platelets are associated with increased risk, which match findings of thrombocytopenia in critical COVID-19 patients.^28^

We recognize that the derivation populations may differ from other populations based both on hospital conditions and inherent demographic differences. An external validation using Seville, Greece, and US populations allows us to assess the broader clinical utility of our findings. The CMR model performs well on these patients, with the strongest performance observed in Seville. Seville consists of a South European population similar to the majority of the derivation cohort. However, it did not face the same capacity challenges as ASST Cremona and HM Hospitals during the study period. Greece had a significantly lower disease spread, resulting in a lower mortality rate compared to the derivation population. Nevertheless, the model yields comparable results in this cohort to the other European hospitals. Hartford has the weakest validation performance, which may suggest inherent differences between Europe and the US in disease dynamics, treatment protocols, or underlying population susceptibility. This attests to the need for training and validation on a diverse set of populations.

We observe that the thresholds needed for obtaining 80% sensitivity differ across the external validation cohorts. When applying the CMR tool to a new hospital, the threshold should be calibrated to the severity of this population. A sample of historical patients at the hospital can be used to validate the model. Using the risk predictions and true outcomes of this sample, various risk thresholds can be evaluated for sensitivity and specificity. Clinicians can determine the relevant threshold for their hospital’s needs. For example, highly constrained systems may employ a higher threshold (lower sensitivity) due to capacity limits, whereas other centers may use this tool as an initial screening tool where sensitivity is required to be very high.

Risk models are most useful when they are readily available for healthcare clinicians. For this reason, a dynamic online application has been created as the interface of the CMR model for use by clinical providers. **Figure 2** provides a visualization of the application that is available at covidanalytics.io/mortality_calculator. After entering a patient’s clinical features, the model returns a predicted mortality risk. It additionally produces a SHAP plot to elucidate the major factors contributing to an individual patient’s risk score. Features in blue decrease risk from the population baseline, whereas features in red increase risk. The contribution is proportional to the width of the feature’s bar. In the example, we see that the patient’s age and oxygen saturation levels increase his risk assessment, but his temperature and glucose lower his risk. The application has been prospectively validated in the emergency room of ASST Cremona to prioritize hospitalizations on higher risk patients.

**Figure 2:**
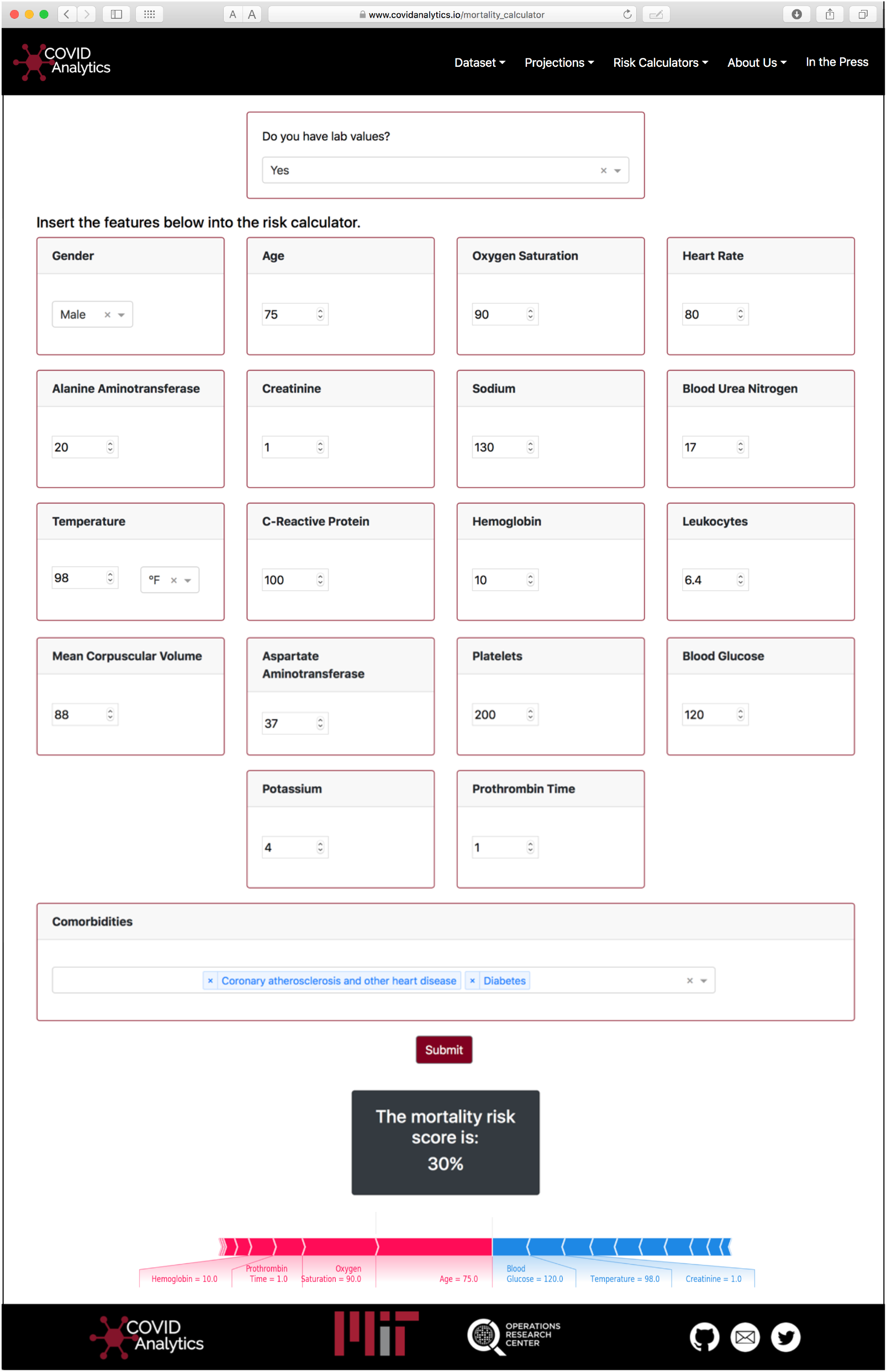
Visualization of the Calculator interface. Using the SHAP package, personalized interpretations of the predicted score are provided to the user.

### Limitations

Limited hospital capacity can impose potential biases in the training population. Only severe patients were able to be treated, particularly in Europe, and some hospitals were forced to turn away patients deemed too critically ill during the peak of the virus. Thus, hospital admissions data may exclude patients on both ends of the acuity spectrum. Additionally, the scarcity of hospital resources may have led patients to receive insufficient care, increasing mortality risk due to lack of treatment. While this warrants further investigation, initial validation results suggest that the CMR tool generalizes well to less congested systems in Greece and the United States.

The differences related to Hartford Hospital might also be related to the timing of the virus. The virus affected Europe before the US. This provided an opportunity to learn from the experience in Europe, which may have resulted in different or more effective treatment decisions as well as governmental policies in the US. This is an opportunity for further study through validation on additional US cohorts.

Our clinical features are limited by the data that was commonly available across all sites in the derivation population. We expect that a more comprehensive set of clinical features such as D-Dimer and IL-6 levels, Body Mass Index, radiographic diagnosis, symptoms, and time elapsed between the disease and treatment onset will yield more accurate results. A broader set of comorbidities, including hypertension, cancer, chronic obstructive pulmonary disease, and others could be included when available. Recent reports on racial disparities and socio-economic determinants of COVID-19 severity^29,30^ could be addressed through the incorporation of additional demographic data and external data sources.

Additionally, there is significant variability in treatment protocols across countries and individual organizations. In future work, we hope to expand the set of captured clinical features and incorporate treatments to disentangle some of the observed heterogeneity in outcomes and clinical characteristics.

## Conclusions

This international study provides a mortality risk calculator of high accuracy for hospitalized patients with confirmed COVID-19. The CMR model validates several reported risk factors and offers insights through a user-friendly interface. Validation on external data shows strong generalization to unseen populations in both Europe and the United States and offers promise for adoption by clinicians as a support tool.

## Data Availability

Electronic Health Record data cannot be shared publicly because it consists of personal information from which it is difficult to guarantee de-identification. As a result, there is a possibility of deductive disclosure of participants and therefore full data access through a public repository is not permitted by the institution that provided us the data. The data and associated documentation from each collaborating institution can only be made available under a new data sharing agreement with which includes: 1) commitment to using the data only for research purposes and not to identify any individual participant; 2) a commitment to securing the data using appropriate measures, and 3) a commitment to destroy or return the data after analyses are complete. Requests can be made to the research team of the corresponding institutions.

## The Hellenic COVID-19 Study Group^†^

Karolina Akinosoglou (University Hospital of Patra), Anastasia Antoniadou (Attikon GH), Katerina Argyraki (Sotiria Thoracic Diseases Hospital of Athens), George N. Dalekos (Department of Medicine and Research Laboratory of Internal Medicine, National Expertise Center of Greece in Autoimmune Liver Diseases, General University Hospital of Larissa), Mina Gaga (Sotiria Thoracic Diseases Hospital of Athens), Nikolaos K. Gatselis (Department of Medicine and Research Laboratory of Internal Medicine, National Expertise Center of Greece in Autoimmune Liver Diseases, General University Hospital of Larissa), Charalambos Gogos (University Hospital of Patra), Ioannis Kalomenidis (Evangelismos Hospital, National and Kapodistrian University of Athens), Eleftheria Kranidioti (Evangelismos Hospital), Lykourgos Kolilekas (Sotiria Thoracic Diseases Hospital of Athens), Eleni Korompoki (Sotiria Thoracic Diseases Hospital of Athens, Department of Clinical Therapeutics, National and Kapodistrian University of Athens), Giota Lourida (Sotiria Thoracic Diseases Hospital of Athens), Evangelia Margellou (Evangelismos Hospital), Georgios Ntaios (Department of Medicine and Research Laboratory of Internal Medicine, National Expertise Center of Greece in Autoimmune Liver Diseases, General University Hospital of Larissa), Periklis Panagopoulos (University Hospital of Alexandroupolis), Angelos Pefanis (Sotiria Thoracic Diseases Hospital of Athens), Vasilis Petrakis (University Hospital of Alexandroupolis), Christos Psarrakis (Attikon GH), Vissaria Sakka (Evangelismos Hospital), Konstantinos Thomas (Attikon GH), Eleftherios Zervas (Sotiria Thoracic Diseases Hospital of Athens).

## Acknowledgements

We would like to thank the HM Hospitals for creating and providing us access to the anonymized clinical data set “COVID Data Saves Lives”. It constitutes a valuable database that is available to the broader research community. In order to gain access to this resource, the reader can contact the following address. We would also like to thank Sophie Testa for the help with the laboratory data collection from ASST Cremona. Moreover, we would like to acknowledge the work of Jeff Mather, Qun Yu, and Lizabeth Roper from the Hartford HealthCare system for their work on data collection. Our team recognizes the contribution and support of José Miguel Cisneros-Herreros, as well as the extensive work of Jesús Moreno-Conde to collect the data from the Hospital Universitario Virgen del Rocío. We would like to thank Álvaro Fernandez Galiana for helping us to get access to the data sources from Spain and reviewing the manuscript. We would also like to thank Aggelos Stefos, Sarah P. Georgiadou, and Anastasia Michail from the Department of Medicine and Research Laboratory of Internal Medicine, National Expertise Center of Greece in Autoimmune Liver Diseases, General University Hospital of Larissa, Larissa, Greece for their help in data collection.

## Other declarations

The investigators were independent from the funders; Luca Mingardi, Agni Orfanoudaki, Bartolomeo Stellato, and Holly Wiberg had full access to the data and can take responsibility for the integrity of the data and the accuracy of the data analysis; Prof. Dimitris Bertsimas affirms that the manuscript is an honest, accurate, and transparent account of the study being reported; that no important aspects of the study have been omitted; and that any discrepancies from the study as planned (and, if relevant, registered) have been explained; Data sharing: no additional data available.

